# Socio-Cultural Factors that Influence Adherence to Mass Drug Administration among Schoolchildren in Schistosomiasis Hotspots along Lake Albert, Hoima District

**DOI:** 10.1101/2025.03.01.25322988

**Authors:** Paskari Odoi, Stella Neema, Fred Bateganya, Birgitte J. Vennervald, Shona Wilson

## Abstract

**Background:** Central to the uptake of praziquantel for control and elimination of schistosomiasis as a public health problem is an understanding of the socio-cultural factors influencing adherence to Mass Drug Administration (MDA) among schoolchildren.

**Methods:** An ethnographic study was conducted to examine socio-cultural factors influencing adherence to MDA among schoolchildren living in the high transmission schistosomiasis area along Lake Albert, Hoima District, Uganda. Nineteen in-depth interviews (IDIs) with schoolchildren, 19 IDIs with parents, 18 Key Informant Interviews, 12 Focus Group Discussions with schoolchildren and unstructured participant observations were conducted. Data was analysed using reflexive thematic analysis. Code reports were inductively generated.

**Results:** Family influence, availability of pre-treatment snacks, increased knowledge of the effectiveness of praziquantel and why someone may or may not experience side effects facilitated praziquantel uptake. This was irrespective of the raised fear of side effects associated with praziquantel, the existence of cultural beliefs and practices and the bitter taste, size, and smell of praziquantel. Children reported that the presence and proximity of sanitation facilities for management of side effects was important. They also suggested lesson free time during MDA would help them manage side-effects.

**Conclusions:** Programmatic factors that could facilitate MDA uptake include continuous health education through integration of schistosomiasis into the school curriculum in transmission hotspots and investment in provision of a pre-treatment snack to reduce side effects. Sanitation as a facilitator of MDA uptake strengthens the argument for continued and expanded integration with the WASH sector for schistosomiasis control.

## Introduction

Human schistosomiasis (bilharzia) is a chronic parasitic disease caused by infection with blood flukes (trematode worms) of the genus *Schistosoma* [1, 2]. The disease is endemic in 78 countries [1], and globally, an estimated 264.3 million individuals needed preventive chemotherapy (PC) in 2022 [3]. Schistosomes are transmitted when freshwater sources are polluted with human excreta (faeces and/or urine) that contain parasite eggs, and individuals are subsequently exposed in the water to the skin-penetrating larval forms (cercariae), which are released after parasite multiplication within freshwater snails. Morbidity is caused by parasite eggs becoming trapped in body tissues and stimulating immune reactions that can progressively damage organs [4].

For the last two decades, schistosomiasis control in endemic areas focused heavily on preventative chemotherapy (PC) through regular Mass Drug Administration (MDA) of the drug praziquantel (PZQ) to school-aged children (SAC). The initial aim was prevention of later-life morbidity by reducing infection levels. This was to be achieved by targeting treatment of 75% of SAC, coupled with wider provision to adults in communities where prevalence in SAC was ≥50% [5]. With the publication of the 2020-2030 roadmap for Neglected Tropical Diseases, the control programme aim has been updated to the elimination of schistosomiasis as a public health problem (<1% of SAC with a heavy infection) and interruption of schistosome transmission in humans in selected countries by 2030 [6]. To date, control has predominately relied entirely on PC with a few exceptions [7]. In sub-Saharan Africa, PC has significantly reduced the prevalence and severity of infection [8]. However, in some areas, often as small as a single village, PC fails to significantly reduce prevalence and intensity of schistosome infections [9]. In such areas, children and adolescents carry the heaviest burden of the infection [10] and, therefore, are a key demographic target for PC.

In Uganda, despite continuous community MDA since 2003 and school MDA since 2004 [11, 12], areas such as Hoima District have maintained persistently high *S. mansoni* infection levels [13]. A recent study among shoreline communities of Lake Albert revealed that although 75% of the respondents acknowledged having previously taken PZQ, only 37% had done so in the past year [13], indicating low programmatic coverage rate. We have previously reported that current inhibiting factors to MDA participation amongst adults in Hoima District include misconceptions, poor community integration and occupational behaviour resulting in non-mobilisation [14]. Similar finding on misconceptions and beliefs were also reported in studies conducted in the early years of PC implementation [15]. However, these earlier studies also attributed poor uptake to fear of side effects, divergence between biomedical and local understandings of schistosomiasis, and inappropriate and inadequate health education, which we found, although present amongst sections of the communities who were hard to reach, were no longer major barriers to MDA uptake amongst adults. Whilst both these studies were conducted in shoreline communities of Lake Albert, their focus on adult household members makes it impossible to identify the views of schoolchildren, the key target demographic of PC. Thus, substantive knowledge gaps remain regarding MDA uptake in this area. We therefore undertook an ethnographic study to examine the socio-cultural factors that influence adherence to MDA among schoolchildren in Hoima District, with the aim of identifying facilitators that could improve control programme coverage and barriers that can be mitigated for in this area and others.

## Methods

### Study area

The study was conducted in the western sub-counties of Kigorobya, Buseruka and Kabaale, Hoima District, around 200 km from Kampala. Agriculture is the major employment above the Lake Albert basin escarpment (Kigorobya and Buseruka sub-counties), while the fishing industry is the major employment near the shores of lake Albert in Kabaale subcounty [23, 24]. For further details, please see [14].

This is a nested study of a larger research programme, FibroScHot, which includes a clinical trial examining the impact of increased PZQ frequency on childhood schistosomiasis. The trial was initiated in February 2020, with the final trial visit conducted in April 2023, coinciding in part with this study. Government MDA using PZQ based on a school delivery system was first initiated in the study area in 2004 [12], and the treatment has been ongoing approximately annually to date.

### Study design

Ethnography studies social interactions, behaviours, and perceptions within groups, teams, organisations, and communities [16]. Engagement with a particular social or cultural group is a distinguishing feature of ethnography [17], with a long-term, holistic, and flexible approach to data collection [18, 19]. The field team, comprising of the first author and a translator, overtly interacted in the daily lives of the study participants for an extended period of ten months from November 2022 to August 2023. This design was employed, first, to allow enriched successive stages of inquiry through the development and refining of tools, techniques, and analysis and second, to allow successive interweaving and triangulation of methods [20]. Triangulation of methods helps to increase the credibility of the findings [21, 22]. Ethnography comprises different ways of eliciting and collecting data; here, we detail the findings from unstructured observation, key informant interviews, in-depth interviews, and focus group discussions.

### Study participants

Participants were pupils of Kaiso and Buhirigi Primary Schools, the participatory schools in the FibroScHot trial. Most study participants speak Alur and a few Lunyoro, the two main local languages. Other participants included parents/caregivers, schoolteachers, health workers, and local government officials. Pupil In-depth Interview (IDI) participants were purposively selected from those proficient and well-informed about schistosomiasis and health, as appropriate for qualitative study design [25–27]. In addition to knowledge and experience, availability, willingness to participate, and the ability to communicate experiences and opinions in an articulate, expressive, and reflective manner were considered [25, 28]. The Focus Group Discussion (FGD) participants were identified, with assistance from the FibroScHot Project coordinators, to gain a wide range of perspectives. Parents/guardians of school pupils were selected as participants in the IDIs. Participation in Key Informant Interviews (KII) depended on roles in the community, including provision of PZQ to the schoolchildren. Although we had planned to interview 76 participants, we only interviewed 68 because saturation, the point in data collection when new data no longer brings additional insights to the research questions [20], was reached.

### Data collection

The four methods of data collection employed are detailed below.

#### Key Informant Interviews

The interviews were conducted at the district, sub-county, and village levels. Participants included district officials, educators, health workers, and community leaders. Eighteen (18) key informants (KIs) out of the planned 20 KIs were interviewed **(see supplementary file one for the key informant interview guide used).**

#### In-depth Interviews

IDIs were conducted with schoolchildren and their parents/guardians (male and female) to understand the socio-cultural factors influencing adherence to MDA among schoolchildren **(see supplementary files two and three for the IDI guides used)**. Children is a contested term, referring to individuals of various ages (see James [29]. In this paper, in line with much other research [30], ‘children’ refers to those under 17, although the minimum age was nine to allow for the ability to articulate. There were 38 IDIs out of the planned 40, 19 in Kaiso and 19 in Buhirigi with schoolchildren and 19 with parents/guardians across both Kaiso and Buhirigi.

#### Focus Group Discussions

FGDs [31, 32] were conducted with children purposively selected from primary grades 4–7 and 9–17 years old, which encompasses the peak age for schistosomiasis infection in Uganda [33]. Each group consisted of six to ten participants. The group category included class. Twelve (12) of the planned 16 FGDs were conducted, with 86 schoolchildren participating in the FGDs **(see supplementary file four for the FGD guide used)**. These interviews were conducted in Alur.

#### Participant observations

Unstructured participant observations were conducted at school, home, and the community to ascertain the sociocultural factors that influence adherence to MDA among schoolchildren. This involved making the participants feel comfortable enough in the researcher’s presence to allow the observation and recording of information about their lives [25].

### Data Management and Analysis

Data was analysed using reflexive thematic analysis following six steps as suggested by Braun and Clarke: 1) familiarisation, 2) coding, 3) generating initial themes, 4) reviewing and developing themes, 5) refining, defining and naming themes, and 6) writing up [34]. Familiarisation involved reading through the transcripts and taking down brief descriptions of potential codes and meanings. This was followed by inductive coding using ATLAS.ti (Version 7). ATLAS.ti was also used to generate coding reports. Each code developed was examined, and what to merge or discard was determined. Initial themes were generated by the 1^st^ author grouping similar codes, followed by an iterative process of regrouping the themes with further review of the codes under each and their meanings. Theme development was a collaborative process to achieve richer interpretations of meaning rather than attempting to reach a consensus of meaning. To a significant extent, participants’ own words are used for the titles of sub-themes to try to provide the reader with a realistic and authentic understanding of the exact words of the participants and their perceptions without being present in the field. Attention was paid to identifying the similarities and differences between participants’ views from the two study sites. All 68 interviews were analysed to avoid missing essential data across participant groups. Evidence from across the identified themes illustrates recurrent patterns that inform on facilitating and inhibiting factors to MDA.

## Results

### Socio-Demographic Characteristics of the Participants

#### In-Depth Interview (IDI)

#### IDI with schoolchildren

The socio-demographic characteristics of nineteen (19) IDI participants showed that 58% of the interviewed participants were females and 53% were from Kaiso Primary School **(Supplementary Table 1).**

#### IDI with parents

The 19 in-depth interview participants showed that 63% of the interviewed participants were females, and 53% had pupils who were studying at Kaiso Primary School **(Supplementary Table 2).**

### Focus Group Discussion (FGD) with Schoolchildren

Six of the 12 FGDs were conducted in Kaiso Primary School and six in Buhirigi Primary School. These interviews were conducted in Alur since all study participants were conversant. By gender, almost all groups involved both males and females **(Supplementary Table 3).**

### Key Informants Interviews (KIIs)

Of the 18 interviews, a quarter were with primary school teachers working in government schools, and more than half were females (**Supplementary Table 4).**

### Facilitating social and cultural factors to adherence to MDA

#### The medicine has helped most of them get rid of worms: Perceived effectiveness of PZQ

The perceived effectiveness of PZQ in the fight against schistosomiasis positively influenced its uptake. According to FGD, schoolchildren and their parents in both study sites, PZQ helped pupils eliminate worms.

> “I took it because I knew the medicine would help me by getting rid of the worms in my stomach.” **(FGD-Female)**

A perceived reduction in the number of smaller and “clean” stomachs was commonly reported by schoolchildren of both study sites, and this motivated future participation. For instance, one participant stated that:

> “My stomach was big, but it started reducing when I started swallowing baya (Alur name for PZQ derived from its first manufacturer Bayer AG and Merck KGaA). That is why when they are giving baya, I also swallow it.” **(IDI-Female Pupil)**

Another participant with a similar sentiment:

> “They say their stomach was swollen, but they swallowed baya and it killed the germ in their stomach, and the germ got finished.” **(FGD-Female)**

A similar finding was also reported during in-depth interviews with parents:

> “They so very much want to take the medicine so that it works on them and they look like the rest of the children.” **(IDI-Male Parent)**

Participants, expressed love for PZQ due to its effectiveness and were okay with its size and taste, thus positively influencing their uptake of PZQ.

> “I like everything about the medicine, even the taste and the size.” (IDI-Female Pupil)

#### I take baya because of the benefits I know I can get from taking it: Participants’ knowledge

Participants in both sites commonly reported that knowledge of PZQ and schistosomiasis was among the factors that influenced their uptake. This knowledge was of the high prevalence of schistosomiasis in their area so that their high risk of contracting schistosomiasis, that PZQ helps get rid of worms and can prevent the stomach from swelling and can be a preventive measure against other symptoms. It was evident across schoolchildren, parents, and key informants that being knowledgeable led to PZQ uptake. In a combined FGD of primary seven and six, a female participant mentioned knowledge of the benefits as a factor.

> “I am not like the rest of the children and do not take that medicine because of the food that is served, but I take it because of the benefits I know I can get from taking it.” **(FGD-Female)**

Other participants reiterated the same:

> “Those children have been told the medicine’s benefits and are aware that taking it will benefit them, so children like that want to participate.” **(IDI-Female Parent)**

Most study participants live near Lake Albert and River Hoimo and have frequent contact with those water bodies. Key informants and other participants reported that their knowledge and perception of being at risk of contracting schistosomiasis led to the uptake of PZQ.

> “When it is time for it to be given, they know themselves that they are at high risk of getting it. They do come, they are cooperative, and we do not have cases reporting with the real disease.” **(KII-Female Health Worker)**

> “I continue to take it so that it kills the worms that are in my stomach, and given the fact that I bathe in the [redacted identifying geographical feature] and drink water from the [redacted identifying geographical feature] somehow, I knew that I had the disease, so it was important for me to take the medicine.” **(IDI-Male Pupil)**

#### Fear of disease and death and a desire for health

Most participants feared getting infected with schistosomiasis, did not want worms to grow, and feared its re-occurrence. Of the twelve FGDs conducted, half featured fear of the disease as a motivator to continue PZQ uptake.

> “They continue because they do not want the worms to continue growing in their stomachs.” **(FGD-Female)**

Others feared falling sick with the disease and the subsequent stigma associated with it.

> “To those who take the tablets……. this is not that they like it so much, but it is because they know that if they do not take them, they will become sick again and other people or children will laugh at them, and maybe to improve their healthy status after seeing the negative effect this disease causes to people who do not take the tablets.” **(KII-Male Health Worker)**

Both schoolchildren and adults acknowledged that the desire to stay healthy motivated their continued participation in MDA.

> “…..because the medicine protects them from diseases and they no longer fall sick like they used to…….” **(FGD-Male)**

> “My children told me that after all the effects of baya, they start feeling better with not much stress. They have started eating well.” **(IDI-Female Parent)**

The findings from IDIs with pupils and their parents revealed that fear of death was among the facilitating factors for PZQ uptake. This fear was sometimes inculcated by treatment providers in order to influence pupils to take PZQ. For example, during one of the IDIs, a pupil stated that:

> “…….there are those who have the disease and they are told if they do not participate then they will die, those also come and participate.” **(IDI-Male Pupil)**

The desire not to die from schistosomiasis and to live long has influenced schoolchildren to participate in MDA.

> “It is because they know that their lives rely on these drugs and without them, they cannot live for so long in health.” **(KII-Male Health Worker)**

#### I had to make them join others and start taking baya: Family influence

Family influence, especially from parents, was central to the uptake of PZQ among schoolchildren. Both parents from the two study sites and schoolchildren from Buhirigi reported this factor during an FGD. In most cases, parents only exert their influence when they start to see the signs and symptoms of schistosomiasis in their children or when pupils have reported refusing PZQ.

> “My child……and some others used not to take, and I realised they were as well developing those signs. I had to make them join others and start taking baya because they were looking miserable and pale.” **(IDI-Female Parent)**

> “Most of the children do not like taking it; for example, my granddaughter herself did not like taking it because she says it does not treat her well at all, but I had to encourage her to take it.” **(IDI-Female Parent)**

On the other hand, some pupils take PZQ because they fear the repercussions from their parents when they learn that they did not participate in MDA. Some of the children reported fearing being caned by their parents for not taking PZQ as a reason for their participation.

> “Some swallow it…because if they take baya home, the parents will cane them.” **(IDI-Male Pupil)**

#### The presence of sanitation facilities at the school

Participants mentioned that having sanitation facilities at school positively influenced their willingness to receive treatment. School-based MDA was preferred in areas with low community latrine coverage. Access to school sanitation facilities was easier for participants, especially during side effects like diarrhea, compared to their communities where such facilities were not readily available. Some areas faced challenges in constructing sanitation facilities due to sandy soil and difficulties in maintaining latrines, especially during the rainy season, leading to collapse.

> “……of course, the soil factor……, now that the rainy season has come, some of the toilets will collapse **(KII-Male Local Government Administrator)**

The presence of sanitation facilities such as latrines and toilets came up, with some of the children preferring to take PZQ at school in case of side effects.

> “…………then there are those who prefer taking baya at school because they know the latrines are near in case they get diarrhoea.” **(FGD-Male)**

A similar finding was reported among FGD participants:

> “Some children do not want to take from home because the latrines are far, and yet sometimes, they get and end up defecating on themselves.” **(FGD-Male)**

### Facilitating project-based factors with potential relevance to MDA delivery

#### At least now, we are remaining with only three girls who are being disturbed: Reduced duration of side effects

The study participants reported that they no longer experience prolonged side effects and some no longer experience any side effects. Participants indicated that it was one of the reasons for schoolchildren’s participation in MDA. The KIIs, especially those directly involved in drug distribution, reported that they have observed that the number of schoolchildren experiencing side effects has reduced:

> “At first, when we gave the first dose, more than 50% of them disturbed us, because children were lining up near the latrine with diarrhoea, so we had to stay so that we monitored them to see if they were improving. We gave them Panadol and Aspirin, but now, at least, we are left with only three girls disturbed by that problem.” **(KII-Female Educator)**

For those who experience side effects, the duration has drastically reduced:

> “We have not got any severe cases where a child stays for almost three days to four days without getting better because after giving the tablets, a child may get the effect for only one day, then the next day in the evening, you find that the child is okay.” **(KII-Female Educator)**

During treatment observations, the participants, especially the pupils under the FibroScHot project, commonly did not experience any side effects.

> “When I take the schistosomiasis medicine, I do not suffer from side effects like the rest of the children. When I take medicine, I can take about three months when the worms have not disturbed me, so because we take medicine a lot, we barely suffer from stomach pains caused by schistosomiasis.” **(FGD-Female)**

In addition, assurance given by drug distributors to alleviate fears regarding why some may experience side effects was also noted:

> “I liked it at the beginning and went to where it was being distributed, and after we all vomited, they said it helps those suffering from worms; now, even if I eat food from the ground, my stomach does not hurt me.” **(IDI-Male Pupil)**

The reduced side effects and increased knowledge are increasing demand for PZQ: “So, there is that increased knowledge, and currently, when you go to the community, especially in areas where specific populations like the cohorts are getting treatment, the general population is demanding that when is ours coming. So, there is this demand for treatment from the general population.” **(KII-Male Local Government Administrator)**

#### The ones who are in the project, before we give them medicine, we first give them something to eat: Pre-treatment snacks

As much as there was evidence of pre-treatment snacks in both the government and project MDA, this was more pronounced in the project. The project had a budget for pre-treatment snacks that included porridge, juice, and bread which were given during treatment visits. During the general MDA by the government, children were reminded to ask their parents to pack them some food, but only a few schoolchildren could afford to come with packed food or buy something during break time. These snacks included biscuits, small cakes, roasted/cooked cassava or sweet potatoes. This was alluded to by the schoolchildren themselves in both study sites and the key informants:

> “………………when they bring baya, they make porridge that brings joy to someone, so someone does not care much about how baya will disturb them but cares about porridge and bread. That makes someone swallow baya.” **(FGD-Male)**

> “For some of them, what makes them happy is what they eat from there. However, now someone is like me. I am taking two cups of porridge, and they say no problem. Give me bread. I want more, they add. So, for them, when they see this person coming, they know. When they call their names, some tell their colleagues we will eat bread for you will not eat.” **(KII-Female Educator)**

Others reported that taking PZQ without a pre-treatment snack tends to make side effects worse for those who experience it.

> “That is true because if you take that medicine without eating, the side effects are even worse.” **(IDI-Male Pupil)**

#### Role of project treatment providers

The care and support provided by the trial team while participants had side effects facilitated PZQ uptake. The most common drug treatments offered by the project was paracetamol for abdominal pain and oral rehydration sachets for those with diarrhoea; but mostly side effects were not treated and the participant reassured instead.

> “But during those days, they would run because they would get some side effects, and then they were not cared for. However, when now they get the drugs, they get some side effects, and they are cared for. So now they have got that curiosity of taking the drug other than the previous days.” **(KII-Female Health Worker)**

### Inhibiting social and cultural factors to adherence to MDA

#### I had a terrible headache, and I also suffered from stomach pain. Perceived fear of side effects

In both study sites, different groups of participants acknowledged fear of side effects as an inhibiting factor in MDA participation. The common side effects that were mentioned included stomach/abdominal pain, stomach ache, diarrhoea, general body weakness, bitter mouth, vomiting, dizziness, sleeping, spitting much saliva, loss of appetite, constipation, frequent visits to the latrine, and high temperature. There were no variations based on whether it was government or project MDA. Below is a snapshot of what schoolchildren reported as side effects that have inhibited their participation in MDA.

> “I felt bad; I suffered from a lot of stomach pain and diarrhoea. I remember my stomach pained me, so I had to sleep under the tree because I wanted to be near the latrine. My stomach kept paining, and after a short while, I started vomiting, and as I was vomiting, I could feel severe heartburn. I kept drinking water and returning to the latrine the whole day,….” **(IDI-Female Pupil)**

The lack of pre-treatment snacks worsened this fear of side effects, especially during the government MDA. This, in combination with the timing of treatment, made some schoolchildren refuse to take PZQ, as they had an empty stomach. These findings were captured in both key informant interviews and school MDA observations.

> “…….for all those years we have been treating, we have relied on the parent’s contribution by children packing food. However, of course, in some communities, there is much scarcity.” **(KII-Male Local Government Administrator)**

> “There was resistance in Buhirigi from the primary seven pupils. Some said it was approaching lunchtime; we cannot take this drug on an empty stomach.” **(Field Notes-Participant Observation)**

#### Schistosomiasis medicine makes you sick immediately: Impact of side effects

The duration of side effects, which some experienced immediately after taking PZQ, inhibited their uptake. A case in point, one of the parents had this to say:

> “The schistosomiasis medicine makes you sick immediately after you swallow it, even though nothing is wrong with you. You immediately start feeling sick when it reaches your stomach, so the child will think they have brought medicine to make them fall sick, so they fear taking it.” **(IDI-Female Parent)**

Due to the impact of side effects, some schoolchildren loose concentration in class while others miss lessons altogether.

> “Sometimes you feel like instead of writing or concentrating in class, you cannot because of too much pain.” **(IDI-Male Pupil)**

It was a common sight during MDA to find some schoolchildren running away while others hide under the desks in an attempt not to swallow PZQ.

> “Sometimes the teacher comes and calls names, then they come out and go to the next class. Some will refuse, and other children run away when they see them coming. Some even hide under the desk……they do not want to swallow baya.” **(IDI-Female Pupil)**

Other schoolchildren, as reported by key informant interviews, rather than refusing PZQ, decide to be absent on days of MDA. Drug distributors do make efforts to follow up, but if they continue to refuse, nothing is done, and they end up not participating in MDA.

> “…………if it is massive (government MDA), we are given two days and those children completely absent themselves, we also leave but if we know maybe the children absented because he is sick or not aware, those we follow them. Nevertheless, those who have refused are those who we leave. We leave them because they have refused.” **(KII-Female Educator)**

On MDA days, most teachers, as reported by participants from Buhirigi, refrain from teaching. When they find children experiencing side effects in class, they move to the next class or return to the staff room.

> “Because of the side effects, they cannot continue with the lesson, and when some teachers come to class and find children suffering from the side effects of the medicine, they do not teach that class. Other children prefer attending class, so they dodge taking the medicine because they know after taking the medicine, they will be unable to attend the class.” **(IDI-Male Pupil)**

#### I do not like that it is bitter and makes your mouth bitter: Fear of size, taste and smell

All participants in this study reported that the bitter taste, size, and smell were among the reasons schoolchildren do not want to participate in MDA. As for the smell, key informants reported that schoolchildren said that PZQ smelled like faeces. Irrespective of the category of FGD and location, they all acknowledged that bitter taste and immense size are inhibiting factors.

> “…..the medicine is bitter, and also they do not allow us to break it into half, and yet it is long, so they end up fearing taking the medicine.” **(IDI-Female Pupil)**

> “They complain it is very bitter. Secondly, they say it smells so badly they compare it to the latrine. Thirdly, it is very big.” **(KII-Female Educator)**

Even when the medicine is broken into two, as observed during MDA for younger children and those who request it be broken, some children still report that PZQ is still long/big.

#### They do not go because they know they do not have it: Feeling healthy

It was also reported that some schoolchildren see themselves as free from schistosomiasis, hence making them not participate in MDA. Perception can be such a powerful tool, and in the context of this study, the fact that some schoolchildren perceive themselves as being free of the disease has negatively affected their participation. For example, during one of the IDIs with a female participant, she stated that:

> “Those children who have schistosomiasis go because they know that baya helps them, but those who do not have schistosomiasis refuse; they do not go because they know they do not have it.” **(IDI-Female Pupil)**

#### If someone knows why they are taking them, you cannot throw those drugs: Ignorance and illiteracy

It was reported that ignorance and illiteracy were among the factors that led to non-participation in MDA in both study sites. Schoolchildren, parents and key informants attributed this to several factors, such as not knowing the benefits of the drug, fear that PZQ would kill them, and lack of prior communication, among others.

> “Some children do not know the benefits of the medicine, but with time, when they learn the benefits, then they will like it.” (**FGD-Male)**

Findings from schoolchildren, parents and key informants indicated that people’s perception of PZQ being able to kill or reduce their life span was among the inhibiting factors to PZQ uptake.

> “……some children do not want to take the medicine because they think the medicine is there to kill them.” **(IDI-Female Pupil)**

As a result of ignorance and illiteracy about PZQ, some of the schoolchildren thought that the medicine did not work, hence losing interest in MDA.

> “They wanted to give me medicine, and I also got medicine, but the germ does not finish still.” **(IDI-Female Pupil)**

It was reported that sometimes schoolchildren come to school without knowing there will be treatment, only to find out when they see the treatment providers’ vehicles in the school compound. Hence, a lack of prior communication was identified.

> “They are not informed. They come to school, and when they see the vehicle coming, then the teachers get the paper moving around reading their names.” **(KII-Female Educator)**

During government MDA, since some children needed persuasion due to the lack of or inadequate sensitisation, some children end up missing treatment since they are unaware of its benefits.

> “These children need to be persuaded……. if the child sees that you are not observing him or her swallowing, that is when they hide. If only they would be sensitised…during massive (government MDA)…. We tell them tomorrow; it is giving drugs.” **(KII-Female Educator)**

#### My aunt refused me to take it, so I have never taken it: Influence of significant others

The influence of significant others was also reported as one of the inhibiting factors of PZQ uptake. Some of the influence came from family members and classmates, among others. A case in point, one child reported that her aunt refused her from participating in MDA.

> “No, I did not because my aunt refused me take it so I have never taken it.” **(FGD-Female)**

Other schoolchildren feared participating in MDA because of the complaints of side effects that they heard from their fellows.

> “Other children hate swallowing the medicine because they hear how their fellow complains about how it has disturbed them.” **(IDI-Male Parent)**

As for the parents, the fact that the medicine makes their children feel like they are dying after taking it is the reason why they are hesitant to allow their children to participate in MDA.

The above influence has made some schoolchildren develop negative attitudes and perceptions about medicine and treatment providers.

> “Children do not trust them because they do not know them, so they sometimes think they bring for them the wrong medicine.” **(IDI-Male Pupil)**

### Inhibiting project-based factors with potential relevance to MDA delivery

#### Every year they take them so for them they don’t like that one: Medicine fatigue

Key informants in this study, especially those directly involved in PZQ distribution, reported medicine fatigue as one of the MDA inhibitors. Schoolchildren under the FibroScHot project are divided into three groups: those who are treated once a year, those who are treated 2 times a year, and those who are treated 4 times a year. As for the rest of the schoolchildren, they are treated under government MDA once a year. As a result, some children have resorted to not participating in MDA.

> “…..you know those children, they are now becoming mature enough; they are taking so many drugs after three months. They have started now thinking…….we are taking drugs so much. So, I would see, some want to hide, some refuse.” **(KII-Male Health Worker)**

## Discussion

We have previously reported on the facilitators and barriers to uptake of preventative chemotherapy amongst adults residing in Hoima District [14]. Here, we reported on the facilitators and barriers amongst SAC, a key demographic target for schistosomiasis control programmes, in the same district. Similar to the findings for adults, the fear of PZQ side effects and dislike of its size and taste are countered by knowledge and lived experience of improved health. There are other commonalities between adults and children, such as family influence, both negative and positive, issues regarding poor sensitisation coverage and knowledge regarding the benefits of treatment. These, in context of other studies, are discussed in detail in [14]. However, three significant findings from the children were not strong themes from the adults, and they are: 1) provision and timing of pre-treatment snack; 2) side effects impacting on lessons/attendance; and 3) the need for sanitation to deal with side effects. Treatment fatigue was also raised by those who provided the PZQ treatment to the children.

Almost all participants raised concerns about the fear of side effects associated with PZQ. This fear of side effects associated with PZQ has been documented in cultures as diverse as Bangladesh [35], the Philippines [36], Zanzibar [37], and mainland Tanzania [38]. Adverse side effects can cause parents and children to hesitate about participating in MDA activities [36, 37]. Crucially, in agreement with Lorenzo *et al.* [36], our results suggest that participants know why they may or may not experience side effects. This knowledge, either of their own or those around them, has greatly helped improve the uptake of PZQ. In addition, those who experienced side effects at the beginning were not deterred from taking PZQ, as found elsewhere in Uganda [39]. Evidence from our study also shows that the lack of pre-treatment snacks during government MDAs increased the fear and occurrence of side effects. Pre-treatment snacks as a motivator for pupils’ participation and a mitigation measure against fear and the occurrence of adverse events has been recommended elsewhere [37, 40–42]. If providing pre-treatment snacks is not possible, then the timing of treatment might be necessary as some pupils refused treatment later in the morning when hungry. Elsewhere in Uganda it was observed that although children and teachers were aware that side effects of PZQ can be mitigated by taking the drug with food, for many children, this was not possible as prevailing poverty resulted in them going to school without a meal, in some cases not having eaten since the night before [40]. The significance of treatment timing in relation to food intake in achieving high coverage has also been reported in the low transmission area of Zanzibar, which is scheduled for interruption of transmission [37], indicating that recognising the importance of snack timing could have a more universal impact, beyond high transmission areas, on control programme uptake.

As much as side effects are not seen as a significant barrier to MDA participation, our findings show an anxiety amongst the children about the transient negative impacts on their learning, and also on teacher behaviour, in relation to continued teaching on the day of treatment. Children reported losing concentration in class and missing lessons altogether. Shared concerns regarding loss of learning, not only among children but also among teachers and community members as a whole have previously been reported [43]. Therefore, to avoid a negative impact on learning, there is a need to look into implementation of innovative strategies, such as lesson-free treatment days in high transmission areas. In addition, inclusion of the longer-term positive effect of preventative chemotherapy on school performance [44], could be emphasised in health education exercises, allowing participants to balance these short-term impacts with longer-term gain. Health education could also help assuage those children who deliberately abscond from school on days of treatment [45].

In the context of schistosomiasis control, WASH infrastructure is usually framed in terms of transmission reduction by reducing both open defecation and the need for water collection from open water that may contain the infective cercaria [4, 46]. However, previous studies conducted among schoolchildren in Jinja District [43] and Koome Island in central Uganda [39] have looked at links between availability of sanitation facilities and uptake of PZQ but failed to identify an association. This was the case in the current study where the presence and nearness of sanitation facilities at school increased compliance. The 2020 Ugandan Water and Environment sector performance report put access to safe sanitation at only seven percent in rural areas [47]. Our finding may therefore be beneficial knowledge for other areas with poor latrine coverage. For example, different highly endemic Ugandan districts employ differing MDA strategies; some, such as Mayuge District, use a community-only approach for drug distribution, while Hoima employs a mixed school and community-based approach. Given the findings here, the combined school and community-based approach may help the MDA treatment coverage rate, not only due to the ease of implementation through school structures [43] but also via a greater uptake where children are re-assured by the presence of school latrines in the event of side-effects.

Finally, and of importance, particularly regarding the WHO guideline of increased treatment to 2x per annum in persistent hotspots [6], medicine fatigue was a reported inhibiting factor to MDA participation by KII participants involved in PZQ distribution. Similar findings have been reported in Côte d’Ivoire [48] and Zanzibar [37]. In Zanzibar ‘treatment fatigue’ was speculated to be due to the population being only marginally infected but repeatedly given medications [37]. Hoima district has already undergone over 15 years of MDA and in the current study, this was compounded by the project’s clinical trial. The Health Belief Model [49] posits that a positive call to action may still occur if the participants’ perceived benefits outweigh their perceived barriers. Other study participants reported they no longer experienced or had reduced side effects and included this reduction as one of their reasons for MDA participation. Moving forward, there is therefore a need for further health education on continued PZQ uptake, if and when treatment frequency is increased.

### Limitations

One limitation of the study is that it only included children from primary four to seven from Kaiso and Buhirigi primary schools; hence, the results may not be generalisable to other schools, or capture the views of children in the younger years of the school-aged demographic. Furthermore, the two schools were not randomly selected but were instead participating in the FibroScHot project. Whilst we cannot adjust our findings for project influence, the comparison between the implementation of treatment via project and via the government MDA has provided vital information. Equally important, the study used a purely qualitative approach, therefore cannot directly link results to coverage rates as would be possible in a mixed-methods study.

### Conclusions and recommendations

We have shown that while side effects per se do not significantly hinder uptake of treatment amongst school-aged children, identified barriers are related to them, and mitigation for these barriers could improve programmatic uptake. We recommend 1) provision of a pre-treatment snack and, where not possible, timing treatment to ensure children are not hungry; 2) that when latrine coverage is low in the community, provision of treatment within the schools where latrines are present is the preferred implementation design, and 3) mitigation against treatment fatigue when incidence of side-effects reduces by integration of continuous health education into the school curriculum. This latter point may be particularly important when treatment frequency is increased in persistent hotspots.

## Supporting information

Focus Group Discussion Guide with the Schoolchildren

Key Informant Interview Guide

In-Depth Interview Guide (parents/guardians)

In-depth Interviews (schoolchildren)

## Acknowledgements

At the time of drafting this manuscript, the first author, Mr. Odoi Paskari, was a postgraduate student in Medical Anthropology at the Department of Sociology and Anthropology, School of Social Sciences, Makerere University. We are grateful for the support from the Department. We thank all members of the wider FibroScHot research team who facilitated the implementation of this anthropological study. The authors acknowledge the contribution from the schoolchildren, parents/guardians, teachers and administrators from Kaiso and Buhirigi primary schools, Hoima District Health Office and the community members of Kabaale, Buseruka and Kigorobya sub counties, who were very supportive and ensured that this study was conducted without any significant challenges. We also acknowledge contributions from Miss. Jovia Adubango, the graduate student who worked alongside the 1^st^ author as an interpreter during data collection.

## Declarations

### Ethical Considerations

The study protocol was approved by the Makerere University School of Social Sciences Research Ethics Committee (MAKSSREC REF 08.2022) and the Uganda National Council for Science and Technology (UNCST REF No SS1463ES). The information leaflets and the consent and assent forms were translated into Alur, the primary local language in Kaiso and Buhirigi. Information leaflets were designed specifically for children and adults and outlined the research aims, methods and timing, and any anticipated benefits, outcomes, risks or costs. Participants were informed during the consent and assent process of confidentiality procedures, and confidentiality was respected during data collection. Consent and assent for each type of instrument were administered separately and before the commencement of the interviews. Before getting children’s assent, we had first to obtain parental or legal guardian consent by signing or thumbprinting consent forms.

## Consent for publication

Not applicable

## Data availability

The data generated or analysed during this study are not publicly available due to the approved study Participant Information Sheets not permitting open access sharing but are available from the corresponding author on reasonable request

## Author Contributions

Conceptualisation (OP, SN, FB); data curation (OP); formal analysis (OP); funding acquisition (SN, SW); investigation (OP); methodology (OP); project administration (OP); supervision (SW, FB); validation (OP, SN, SW); writing – original draft (OP); writing – review and editing (SN, FB, BJV, SW).

