## Supplementary material for "Socio-Cultural Factors that Influence Adherence to Mass Drug Administration among Schoolchildren in Schistosomiasis Hotspots along Lake Albert, Hoima District": Focus Group Discussion Guide with the Schoolchildren

**Supplementary File Four: Focus Group Discussion Guide with the Schoolchildren**

**A Form for social and demographic characteristics of FGD participants**

| **FGD_NO** | **Social category: (e.g. An FGD schoolchildren** |
| --- | --- |
| **Name of group (e.g. Male FGD)** |  |

| **DATE** |  | **District** |
| --- | --- | --- |

| **Place of discussion** | (e.g. school) |
| --- | --- |

| **Moderator**  **Recorder/note-taker** | **Name or Code** |
| --- | --- |

| **Recording system (to be linked to audio file)** | (e.g. digital recording + notes taking) |
| --- | --- |

| **Time** | **Start:** | **Finish:** | **Duration:** |
| --- | --- | --- | --- |

Participant identifier

| **S/No** | **Participants number** (e.g. 01) | **Sex**  M/F | **Age** | **Education** | **Religion** | **Ethnic group** | **Ever had Bilharzia** |
| --- | --- | --- | --- | --- | --- | --- | --- |
| **1.** |  |  |  |  |  |  |  |
| **2.** |  |  |  |  |  |  |  |
| **3.** |  |  |  |  |  |  |  |
| **4.** |  |  |  |  |  |  |  |
| **5.** |  |  |  |  |  |  |  |
| **6.** |  |  |  |  |  |  |  |
| **7.** |  |  |  |  |  |  |  |
| **8.** |  |  |  |  |  |  |  |

**Purpose:**

- To establish the sociocultural factors that influence adherence to MDA among schoolchildren
- To assess the interaction between Schoolchildren and the providers of treatment during MDA
- To document the daily lived experiences of schoolchildren that shape adherence to MDA

**Key Domains and Questions**

**a) Understandings of illness:**

We are interested to learn how schoolchildren in this school understand illness in general - and bilharzia (use local name) in Particular.

1) In your view where does illness come from?
2) What are the main sources of illness that can make a person unwell in this community?
3) How do schoolchildren come into contact with Bilharzia?
4) What are local terminologies used to describe Bilharzia? Why those terms?

**b) The experience of bilharzia**

5) Is bilharzia common in this place?
6) When a person has bilharzia, how does he/she feel? What are the symptoms?

*Probe: are these serious symptoms or something one just lives with?*

7) How do these symptoms affect everyday life?

*Probe: do these symptoms ever persist or return – and how does that make a person feel – what does it make them do?*

8) Is there any stigma associated with these particular symptoms/bilharzia? (how do people respond to your symptom experience?)

*(a) Probe: If there is stigma - Why are these symptoms stigmatised?*

*(b) Probe: If there is stigma - How is stigma expressed?*

*(c) Probe: If there is Stigma - How is it experienced by a person with the symptoms – and how does it affect what they do about their symptoms?*

*(d) Probe: Are things the same or different for different groups (boys/girls).*

**c) Pathways to care**

We are interested to learn what schoolchildren do when they become ill with bilharzia.

9) When schoolchildren in this community get bilharzia what do they do? Where do they get treatment? probe for biomedical such as hospitals (levels), clinics, traditional/folk-herbs, witch doctors/traditional healers etc) Which treatment? *(probe for praziquantel, herbs, concoctions etc)*
10) When they fail to get better what else do they do?

*Probes: alternative care or traditional care (herbal etc.)? When do they do that and why?*

11) Kindly tell us more about situations when schoolchildren go for MDA? What do they do there and why?
12) Please tell us whether you took MDA in the last treatment and why? And whether you are willing to take the drugs in the future? *Explain*

**d) Risk factors for bilharzia**

13) What are those things that can help the spread of bilharzia?
14) Why do some schoolchildren in this community get bilharzia while others do not get it?
15) What makes some Schoolchildren prone to getting bilharzia?

*Probe for Gender, age, education, residence, and ethnic difference*s

**e) Inhibiting and facilitating factors in the uptake and adherence to praziquantel MDA among schoolchildren**

16) Schoolchildren suffering from bilharzia are given a medicine called praziquantel. Tell me about it. What do schoolchildren who have taken praziquantel talk about it?

- What do they like about the medicine? Why do they want to continue taking the drug? (motivating factors)
- Why do some schoolchildren not want to take the medicine? Who are those children who don’t want to take medicine? What don’t they like about the medicine? *Probe for the barriers- taste, size, colour, the person administering, time and place etc*
- What do you think should be done to improve the uptake of the medicine?
- What should be done to improve adherence to MDAs? Who should do it?

***We are coming to the end of our discussion. Is there anything else you would want to share with us about what we have been discussing?***

***Do you have any questions you would like to ask us – we will do our best to answer them***

***Thank you so much for your time and insights.***
