## Supplementary material for "Socio-Cultural Factors that Influence Adherence to Mass Drug Administration among Schoolchildren in Schistosomiasis Hotspots along Lake Albert, Hoima District": Key Informant Interview Guide

**Supplementary File One: Key Informant Interview Guide**

A. **Background information**

| ***No*** | ***Background information*** | ***Response*** |
| --- | --- | --- |
| a. | Date of interview |  |
| b. | Level (District, sub-county, community, school) |  |
| c. | Type of organization working for (e.g. government, private, church, other) |  |
| d. | Sex of informant |  |
| e. | Informant’s highest level of education (e.g. Certificate, diploma, degree, post-graduate degree) |  |
| f. | Informant’s designation (e.g. Medical Officer, Clinical Officer, Teacher etc.) |  |
| g. | Interviewer’s code |  |

B. **Purpose**

- To establish the sociocultural factors that influence adherence to MDA among schoolchildren;
- To assess the interaction between schoolchildren and the providers of treatment during MDA;
- To document the daily lived experiences of schoolchildren that shape adherence to MDA.

**Key Domains and Questions**

**A) Understandings of illness:**

1) We are interested to learn how schoolchildren in this area understand illness in general - and Bilharzia (use local name) in Particular.
i. Let us start by asking in very general terms – in your experience and opinion, what do people think causes bilharzia?
ii. Is bilharzia common in this community/district/sub-county?
iii. Is bilharzia more common in some schoolchildren than others? Please explain
iv. Is there any stigma associated with particular symptoms/bilharzia? Why are these symptoms stigmatised? How is stigma expressed? How do schoolchildren deal with the stigma?

**B) Pathways to care**

2). In your experience and opinion, when schoolchildren in this community get bilharzia what do they do?

i. Where do they get treatment? (probe for different sources of medication-biomedical, traditional/alternative sources) Which treatment? (probe for: praziquantel, herbs, concoctions)
ii. When they fail to get better what else do they do?

*Probes: alternative care or traditional care (herbal etc.)? When do they do that and why?*

**C) Risk factors for bilharzia**

3) Several things can help the spread of bilharzia.

i. Please explain what you think they are in this community?
ii. Why do some schoolchildren in this community get bilharzia while others do not get it?
iii. What are those factors that make some schoolchildren prone to getting bilharzia?

*Probe for Gender, age, education, residence, ethnic differences, etc)*

**D) Inhibiting and facilitating factors in the uptake of praziquantel treatment among Schoolchildren**

(4) Schoolchildren suffering from Bilharzia are given a medicine called Praziquantel.

i. What do schoolchildren who have taken praziquantel talk about it? Any side effects?
ii. What do schoolchildren like about the medicine?
iii. Why do schoolchildren want to continue taking the drug? (motivating factors)
iv. Why do some schoolchildren not want to take the medicine?
v. What don’t schoolchildren like about the medicine? *Probe for the barriers- taste etc*
vi. What are the effects of bilharzia: *probe at the individual, household, and community?*
vii. In your opinion what do you think should be done to improve the uptake of the medicine among schoolchildren?

**E) Inhibiting and facilitating factors in the adherence to Mass Drug Administration**

(5) The bilharzia medicine (Praziquantel) is distributed to schoolchildren during the mass drug administration (MDA). We want to know why some schoolchildren like to participate while others do not.

i. Why do schoolchildren want to participate in MDA in this community? Who are those schoolchildren who usually participate?
ii. Why do some schoolchildren not want to participate in MDA in this community?
iii. Who are those schoolchildren who usually don’t want to participate?
iv. What should be done to improve adherence to MDAs among schoolchildren? Who should do it?
v. As a key stakeholder, what final suggestion do you have to eliminate bilharzia in your district/community?

***We are coming to the end of our discussion. Is there anything else you would want to share with us about what we have been discussing?***
