## Supplementary material for "Socio-Cultural Factors that Influence Adherence to Mass Drug Administration among Schoolchildren in Schistosomiasis Hotspots along Lake Albert, Hoima District": In-Depth Interview Guide (parents/guardians)

**Supplementary Files Three: In-Depth Interview Guide (parents/guardians)**

***Target****: Parents/guardians of the selected schoolchildren who are in the selected primary schools*

***Aim:*** *To document the daily lived experiences of your schoolchild/children that shape adherence to MDA and the pathways to care.*

**Background Information**

| ***No*** | ***Background information*** | ***Responses*** |
| --- | --- | --- |
| a. | Date of interview |  |
| b. | Sub-county |  |
| c. | Village |  |
| d. | Gender |  |
| e. | Age |  |
| f. | Marital status |  |
| g. | Highest level of education |  |
| h. | What do you do every day? |  |
| i. | Ethnicity |  |
| j. | Religion |  |
| k. | Ever heard bilharzia? |  |

**Key Domains and Questions**

**A) Understandings of illness:**

1. We are going to talk about bilharzia (use the local name).
i. Let us start by asking in very general terms – what is bilharzia?
ii. In your experience and opinion, what do you think causes bilharzia among School children?
iii. Is bilharzia common in this community?
iv. Is bilharzia more common in some schoolchildren than others? Please explain
v. When do schoolchildren get bilharzia? How do they feel?
vi. How do you think schoolchildren contract it?
vii. What are the symptoms that affect schoolchildren the most?
viii. Did people stigmatize schoolchildren when they contracted bilharzia? *Explain*
ix. How do schoolchildren deal with the stigma?

**B) Pathways to care**

2. When schoolchildren get bilharzia, what do you do?

i. Where do you take them for treatment? (probe for different sources of medication-biomedical, traditional/alternative sources) Which treatment do they get? (probe for: praziquantel, herbs, concoctions)
ii. When schoolchildren failed to get better what else do you do?

*Probes: alternative care or traditional care (herbal etc.)? why?*

**C) Risk factors for Bilharzi**a **among Schoolchildren**

3. Several things can help the spread of bilharzia among schoolchildren.

i. Please explain what you think these things are that help the spread of bilharzia in this community.
ii. Why do some schoolchildren in this community get bilharzia while others do not get it?
iii. Specifically, for your schoolchild/children, why do you think he/she or they got infected with bilharzia?
iv. What are those factors that make some schoolchildren prone to getting bilharzia? (*Probe for Gender, age, level of education, location, ethnic differences etc)*
v. What are the effects of bilharzia? *(Probe at the individual, household, and community)*

**D) Inhibiting and facilitating factors in the uptake of praziquantel treatment among schoolchildren**

4. Schoolchildren suffering from bilharzia are given a medicine called praziquantel.

a) Were they given this medicine? If yes, tell me more about it. Are there any side effects?
b) What do you think they liked about the medicine? Did they talk about the side effects also?
c) Why did they continue taking the drug? (motivating factors)
d) Why do some schoolchildren not want to take the medicine? (probe for category of schoolchildren)
e) Did schoolchildren at any time think of not taking the medicine?
f) What don’t schoolchildren like about medicine? *Probe for the barriers- taste etc*
g) What can prevent schoolchildren from taking the medicine
h) What are the effects of not taking praziquantel medicine? *probe at the individual, household, and community?*
i) In your opinion what do you think should be done to improve the uptake of the medicine

**E) Inhibiting and facilitating factors in the adherence to Mass Drug Administration**

5. The bilharzia medicine (praziquantel) is distributed to schoolchildren during the mass drug administration (MDA). We want to know why some schoolchildren like to participate while others do not.

i. Why do schoolchildren want to participate in MDA in this community? Who are those who usually participate?
ii. Why do some schoolchildren not want to participate in MDA in this community?
iii. Who are those who usually don’t want to participate?
iv. Did your schoolchild/children participate in MDA? If yes, why?
v. What should be done to improve adherence to MDAs? Who should do it?
vi. What final suggestion do you have to help eliminate bilharzia in your community?

***We are coming to the end of our discussion. Is there anything else you would want to share with us about what we have been discussing?***

***Thank you very much for your time and ideas***
