## Supplementary material for "Socio-Cultural Factors that Influence Adherence to Mass Drug Administration among Schoolchildren in Schistosomiasis Hotspots along Lake Albert, Hoima District": In-depth Interviews (schoolchildren)

**Supplementary Files Two: In-depth Interviews (schoolchildren)**

***Target****: Schoolchildren who are in the selected primary schools*

***Aim:*** *To document the daily lived experiences of schoolchildren that shape adherence to MDA and the pathways to care*

**Background Information**

| **No** | **Background characteristics** | **Responses** |
| --- | --- | --- |
| a. | Date of interview |  |
| b. | Sub-county |  |
| c. | Village |  |
| d. | Whom do you live with |  |
| e. | Gender |  |
| f. | Age |  |
| g. | Primary level |  |
| h. | Ethnicity |  |
| i. | Religion |  |
| j. | Ever hard Bilharzia? Family member ever had bilharzia |  |

**Key Domains and Questions**

**A) Understandings of illness:**

1. We are going to talk about bilharzia (use the local name).
i. Let us start by asking in very general terms what bilharzia is.
ii. In your experience and opinion, how can a schoolchild get bilharzia?
iii. How would you know that one has bilharzia?
iv. Is bilharzia common in this community?
v. Is bilharzia more common in some schoolchildren than others? Please explain
vi. Have you ever gotten bilharzia? If yes, how did you feel?
vii. How do you think you contracted it?
viii. What were the symptoms that affected you most?
ix. Did other children label you? Explain
x. How did you deal with the stigma?
B) **Pathways to care among schoolchildren**
2. When you got bilharzia, what did you do?
i. If you got treatment, where did you go for treatment?
ii. Which treatment did you get?
iii. Did the treatment help you to get better?
iv. If you failed to get better, what else do you do?

P*robes: alternative care or traditional care (herbal etc.)? why?*

**C) Risk factors for Bilharzia** **among Schoolchildren**

3. Several things can help the spread of bilharzia.

i. Please explain what you think they are in this community?
ii. Why do some schoolchildren in this community get bilharzia while others do not get it?
iii. Specifically, for you, why do you think you got infected with bilharzia?
iv. What are those factors that make some schoolchildren prone to getting bilharzia? (*Probe for Gender, Age, Location, Ethnic differences, etc.)*
v. What are the effects of Bilharzia: *probe at individual, household, community?*

**D) Inhibiting and facilitating factors in the uptake of praziquantel treatment among School children**

4. Schoolchildren suffering from bilharzia are given a medicine called praziquantel during MDA.

i. Were you given this medicine? Tell me more about the medicine? Any side effects?
ii. What did you like about the medicine?
iii. Why did you continue taking the drug? (motivating factors)
iv. Why do some schoolchildren not want to take the medicine? (probe for category of schoolchildren)
v. Did you at any time think of not taking the medicine? *Explain*
vi. What don’t you like about the medicine? *Probe for the barriers- taste etc*
vii. What can prevent you from taking the medicine?
viii. What effects will it have if you do not take praziquantel medicine? (probe for individual, household, community)
ix. In your opinion what do you think should be done to improve the uptake of the medicine

**E) Inhibiting and facilitating factors in the adherence to Mass Drug Administration among Schoolchildren**

5. The bilharzia medicine (praziquantel) is distributed to schoolchildren during the mass drug administration (MDA). We want to know why some schoolchildren like to participate while others do not.

i. Why do schoolchildren want to participate in MDA in this community? Who usually participates?
ii. Why do some schoolchildren not want to participate in MDA in this community?
iii. Who are those schoolchildren who usually don’t want to participate?
iv. Did you participate in the last MDA? If yes, explain why.
v. What should be done to improve adherence to MDAs? Who should do it?
vi. What final suggestion do you have to help eliminate bilharzia in your community?

***We are coming to the end of our discussion. Is there anything else you would want to share with us about what we have been discussing?***

***Thank you very much for your time and ideas***
