## Supplementary Table 1-4 for "Socio-Cultural Factors that Influence Adherence to Mass Drug Administration among Schoolchildren in Schistosomiasis Hotspots along Lake Albert, Hoima District"

| **Gender** | **N (%)** |
| --- | --- |
| Male | 8 (42%) |
| Female | 11 (58%) |
| **School** |  |
| Kaiso Primary School | 10 (53%) |
| Buhirigi Primary School | 9 (47%) |
| **Heard of Bilharzia** |  |
| All | 19 (100%) |

| **Gender** | **N (%)** |
| --- | --- |
| Male | 7 (37%) |
| Female | 12 (63%) |
| **Site** |  |
| Kaiso primary school | 10 (53%) |
| Buhirigi primary school | 9 (47%) |

discussion participants (schoolchildren)

| **FGD** | **School** | **Number of**  **participants** | **Gender** |
| --- | --- | --- | --- |
| 1 | Buhirigi | 7 | Male &  Female |
| 2 | Buhirigi | 7 | Male &  Female |
| 3 | Buhirigi | 8 | Male &  Female |
| 4 | Buhirigi | 7 | Male &  Female |
| 5 | Buhirigi | 10 | Male &  Female |
| 6 | Buhirigi | 8 | Male &  Female |
| 7 | Kaiso | 6 | Female |
| 8 | Kaiso | 6 | Male &  Female |
| 9 | Kaiso | 6 | Female |
| 10 | Kaiso | 6 | Male &  Female |
| 11 | Kaiso | 7 | Male &  Female |
| 12 | Kaiso | 8 | Male &  Female |

| **Gender** | **N (%)** |
| --- | --- |
| Male | 8 (44%) |
| Female | 10 (56%) |
| **Designation** |  |
| Health worker | 9 (50%) |
| Educator | 6 (33%) |
| Government | 3 (17%) |
